# Seroprevalence and risk factor investigation for the exposure of *Toxoplasma gondii* among veterinary personnel in Punjab, India

**DOI:** 10.1101/2020.10.20.20216556

**Authors:** R. Thakur, R. Sharma, R.S. Aulakh, J.P.S. Gill, B.B. Singh

## Abstract

*Toxoplasma gondii*, a globally important food borne zoonotic parasite, infects most of the warm blooded animals as well as people. One third of worlds population has been exposed to *T. gondii* at least once in their lifetime. Veterinarians and para vets are considered at risk of *T. gondii* exposure. As far as we are aware, occupational exposure of *T. gondii* has not been systematically explored from north India. We determined the seroprevalence of *T. gondii* in veterinary personnel and investigated associated risk factors in Punjab, India. Two hundred and five blood samples collected from veterinary personnel were tested for the presence of *Toxoplasma* IgG and IgM antibodies using ELISA. The apparent and true seroprevalence of *T. gondii* with 95% confidence interval (CI) were estimated. Information about participant demographics, and possible routes of exposure was collected using a self completed questionnaire at the time of blood collection. A veterinary person was considered *Toxoplasma* seropositive using a combination of tests in parallel, i.e. if it was positive in either IgG or IgM ELISA. A mixed effects logistic regression model was constructed to evaluate the association of demography, occupational and non-occupational factors with *Toxoplasma* seropositive status. The apparent and estimated true seroprevalence of *T. gondii* antibodies using *Toxoplasma* IgG ELISA was found to be 8.78% (95% CI 5.63% - 13.45%) and 7.36% (95% CI 4.04% - 12.29%), respectively. The apparent and estimated true seroprevalence using *Toxoplasma* IgM ELISA was found to be 0.49% (95% CI inestimable - 2.71%) and 0.51% (95% CI inestimable - 2.83%), respectively. After adjusting for other variables in the final model, consuming mutton and owning a cat were associated with large odds of being *Toxoplasma* seropositive. We report that occupational risk factors are not associated with *Toxoplasma* seropositivity in veterinary personnel in Punjab, India. The seroprevalence of toxoplasmosis in veterinary personnel is comparatively low and occupational exposure in veterinary personnel does not enhance risk of getting infected with *T. gondii* in Punjab, India.

## Introduction

*Toxoplasma gondii* is an intracellular obligate apicomplexan parasite having a broad geographic and host range. This parasite is of worldwide distribution and can infect most of the warm-blooded animals including humans, mammals, and birds (Robert-Gangneux and Dardé, 2012). *Toxoplasma gondii* is the single species in the genus *Toxoplasma*, although genetic diversity within the species has been reported (Ajzenberg et al., 2002). Three well recognized parasitic stages tachyzoite, bradyzoite (contained in cyst) and oocyst have been identified. Cats act as definitive host and shed oocysts in their faeces (Frenkel et al., 1970; Dubey and Frenkel, 1972). Warm blooded animals become infected after ingesting the infective oocysts (Black and Boothroyd, 2000). Human infection could occur through consuming either food or water contaminated with oocysts or infected meat containing tissue cysts (Hill and Dubey, 2002). Human infection can be acquired postnatally or congenitally as well.

*Toxoplasma gondii* is a well-recognized zoonotic parasite and has been reported to be affect approximately 25% to 30% of the global human population (Flegr et al., 2014; Hussain et al., 2017). Further, HIV infected people have a high prevalence rate of toxoplasmosis (Wang et al., 2017). However, the prevalence rates vary between countries or within communities (Montoya and Liesenfeld, 2004; Pappas et al., 2009). Environment as well as livestock play an important role for parasite prevalence in humans (Robert-Gangneux and Dardé, 2012). Globally, 190,100 annual cases of congenital toxoplasmosis have been reported, leading to 1.20 million Disability-adjusted life years (DALYs) and 1.5 cases of congenital toxoplasmosis per 1000 live births (Torgerson and Mastroiacovo, 2013).

Infections in immunocompetent hosts could range from asymptomatic to symptoms such as lymphadenitis, myocarditis or chorioretinitis (Weiss and Dubey, 2009). Congenitally acquired infections are more severe during the first trimester than in the late pregnancy. Congenital toxoplasmosis could result in slightly diminished vision to chorioretinitis, hydrocephalus, convulsions and intra-cerebral calcification (Hill and Dubey, 2002). The disease manifestation in humans could range from asymptomatic to life-threatening in individuals with impaired immune system (Weiss and Dubey, 2009). In immuno-compromised individuals, the disease leads to encephalitis and is one of major cause of death in AIDS patients (Weiss and Dubey, 2009). Studies suggest that 10 to 20% of *T. gondii* infections could result in symptomatic manifestations (Remington et al., 1960).

Besides being an important food-borne pathogen, *T. gondii* is believed to be an occupational hazard among veterinary personnel, laboratory workers, slaughterhouse workers, military workers, forestry workers, and workers exposed to unwashed raw fruits and vegetables, sewage, and soil (Zimmermann, 1976; Swai and Schoonman, 2009; Alvarado-Esquivel et al., 2010; Alvarado-Esquivel et al., 2011; Gómez-Marín et al., 2012; Sang-Eun et al., 2014). Veterinary personnel are considered at a higher risk of getting infected in clinical settings (Tizard and Caoili, 1976) and not adopting personnel protective measures (wearing gloves, lab coats, coveralls) are the potential risk factors for toxoplasmosis.

Toxoplasmosis is endemic in India. Several studies have documented *Toxoplasma* exposure in humans (Dhumne et al., 2007; Singh et al., 2014) and animal hosts (Singh et al., 2010; Thakur et al., 2019). Jani and colleagues (RG et al., 2006) reported 15% (n=60) seroprevalence of toxoplasmosis in zoo workers in western India; higher sero-prevalence was seen in zoo attendants working with feline animals followed by reptiles (RG et al., 2006).

As far as we are aware, occupational exposure of *T. gondii* has not been systematically explored from north India. In view of this, the current study was planned to determine seroprevalence and risk factor investigation for the exposure of *T. gondii* in veterinary personnel in Punjab, India.

## Methods

### Ethics statement

The current study was approved by Institutional Ethics Committee, Dayanand Medical College & Hospital, Ludhiana, Punjab (Ethics approval number: DMCH/ R&D/2018/683)

### Study area

Punjab (Latitude of 30° 4′ ;N and Longitude 75° 5′ E), a north Indian state having 22 districts, is home to 27.7 million people(COI, 2011). The state has a livestock population of 8.12 million primarily consisting of cattle (2.43 million) and buffalo (5.16 million)(BAHS, 2014). Other important livestock species reared in the state include sheep (n=128534), goat (n=327272) and pigs (n=32,221) (BAHS, 2014). No objective information on the number of cats is available, although both stray and pet cats reside in the state.

### Target and study population

The target population consisted of veterinary personnel (Professor/academicians, veterinary doctors, veterinary pharmacists, animal attendants) and students working with livestock in Punjab. As per official data, there were 772 veterinarians, 1762 veterinary pharmacists and 2316 animal attendants serving in the Department of Animal Husbandry, Dairying and Fisheries (DAHDF), Government of Punjab(DAHP, 2012). In addition, there are approximately 500 veterinary students (4^th^ and final year of undergraduate course, Masters and PhD students) and clinical veterinary complex faculty (Pers commun J. Singh) comprising a total population of 5350 veterinary personnel in Punjab.

The study population consisted of all the veterinary personnel working in the DAHDF in Ludhiana district of the state, fourth and final year undergraduate, masters and PhD students enrolled in College of Veterinary Sciences and clinical veterinary complex faculty, Guru Angad Dev Veterinary & Animal Sciences University, Ludhiana.

### Sample size

Assuming a seroprevalence of 14% as reported for the general public (Dhumne et al., 2007) and a population size of 600, the study would have required a sample size of 142 for estimating the prevalence with 5% absolute precision and 95% confidence (NK and MS, 2014). After considering potential clustering at the subdistrict level using a design effect of 1.5, a sample size 213 was required. However, with no prior information on the disease prevalence and level of clustering, all those subjects who consented to participate were enrolled.

### Enrolment

A cover letter and participant information statement briefly describing objectives of the study were provided to the veterinary personnel working in district Ludhiana. Veterinary personnel were contacted during alumni meet and during their monthly meetings. A formal letter was displayed on notice board of the college of veterinary science as well as student hostel(s) explaining objectives of this study and requesting the students to participate in this study. Lastly, academicians working in clinical veterinary complex, GADVASU were personally invited. Those who consented to participate were enrolled in the study.

### Blood Sampling

Five ml of blood was collected from each participant in 2017-18. Serum was separated from clotted blood by centrifuging at 1,200 rpm for 10 min and stored in screw–caped sterilized vials at –20°C until further testing.

### Questionnaire design

A detailed questionnaire was designed to collect information on demography and putative risk factors associated with *T. gondii* infection in veterinary personnel. Information on demography, risk of occupational exposure, past incidence, perception and practices associated with toxoplasmosis were included. The questionnaire was available in two languages: English and Punjabi (local language of the state).

### Laboratory testing

The commercially available NovaLisa™ *T. gondii* IgG and IgM capture-ELISA kits manufactured by NovaLisa™, GenWay Biotech, Inc. 6777 Nancy Ridge Drive San Diego, CA were procured. The IgG and IgM ELISAs were carried out as per manufacturer’s instructions.

The results of IgG ELISA were quantified in IU/ml. The mean absorbance values (on the vertical y–axis) of the four standards A, B, C and D (provided in IgG ELISA kit) were plotted against their corresponding concentrations (0, 50, 100 and 200 IU/ml) (on the horizontal x– axis) and a calibration curve was drawn. Linear regression was used for each test serum sample for obtaining quantitative results of IgG ELISA in IU/ml. Results were interpreted as (a) reactive (> 35 IU/ml), (b) grey zone/equivocal (30 – 35 IU/ml) and (c) non–reactive (< 30 IU/ml).

The IgM ELISA results were interpreted as per the formula provided in manufacturer’s guidelines.

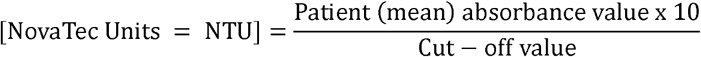

Test serum samples having a cut off value greater than 11 NTU were considered as positive, 9–11 NTU as equivocal and less than 9 NTU were considered negative. The samples within equivocal zones were reported as inconclusive cases as antibodies against pathogen could not be clearly detected.

### Outcome variable

A human subject was considered as *Toxoplasma* seropositive by using the combination of tests in parallel, i.e. if person was seropositive in either *Toxoplasma* IgG or IgM ELISA (Table 1).

**Table 1.**
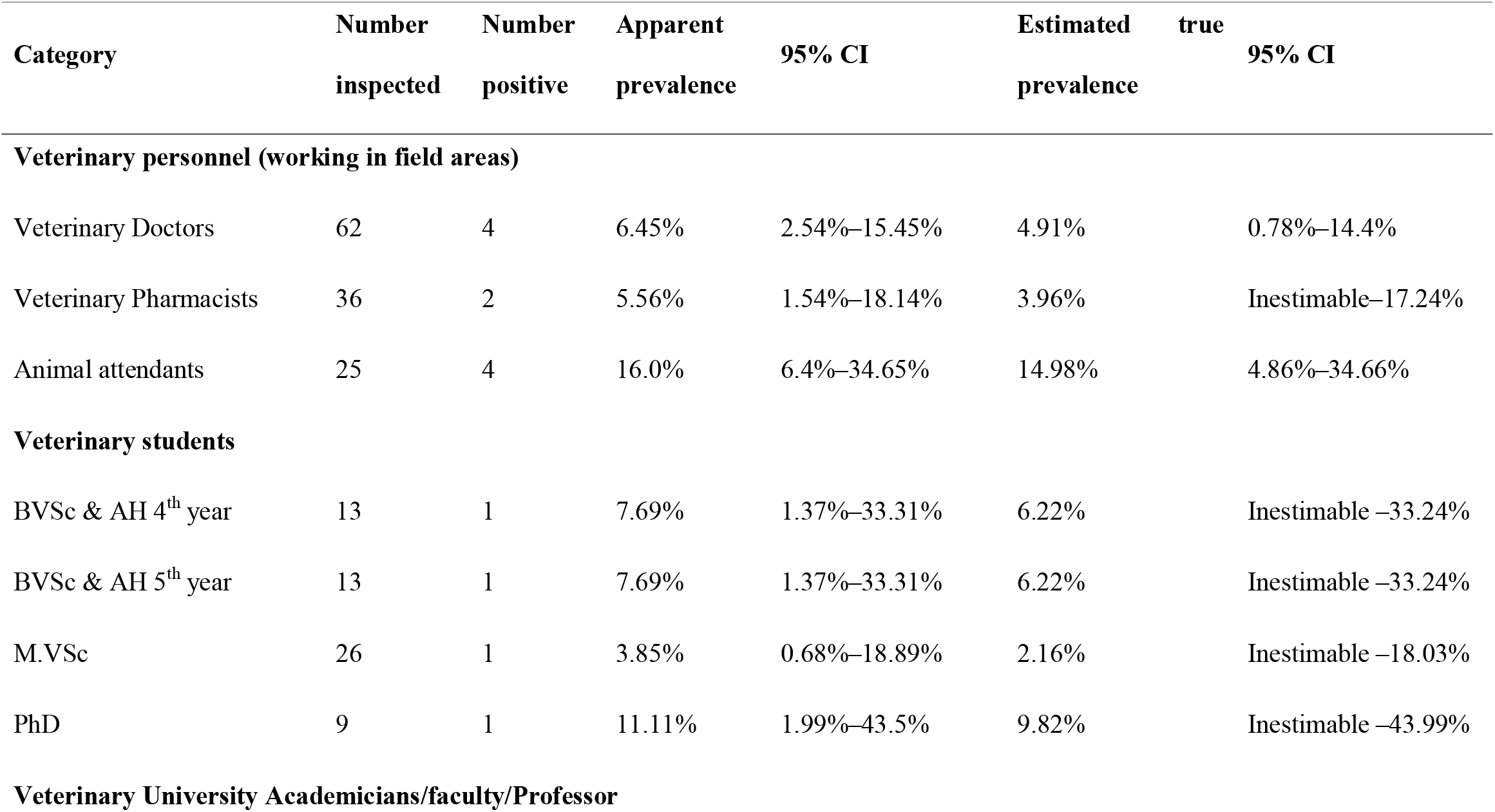
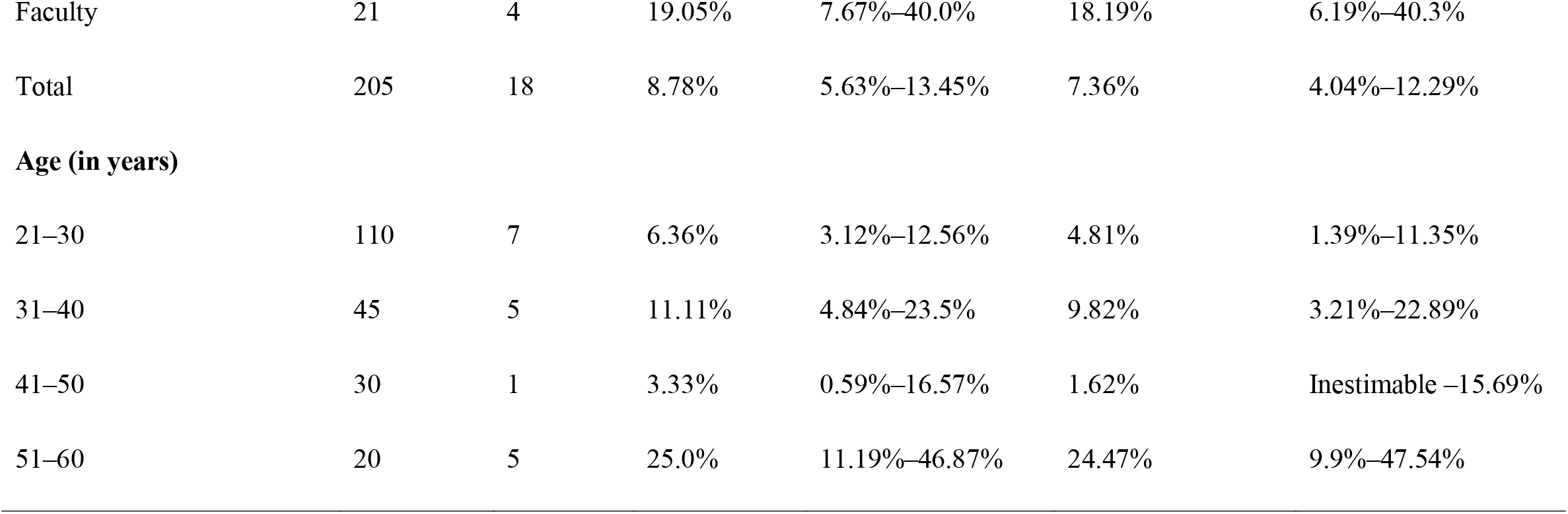
Apparent and estimated true seroprevalence of *T. gondii* antibodies using IgG ELISA in veterinary personnel and students in Punjab, India

### Explanatory variables

A total of thirty–one explanatory variables were considered in this study (Supplementary appendix): demographic (eight variables), non-occupational (eleven variables) and occupational risk exposure variables (twelve variables). Occupational risk exposure variables included animal cases handled (species wise) and personal protective equipment (PPE) use. Information on non-occupational exposure, for example having cat as a pet, gardening practices, meat preferences and habit of consuming raw vegetables and fruits, were also used.

### Statistical analyses

The apparent and true seroprevalence of toxoplasmosis with 95% confidence interval (CI) were determined using the given diagnostic sensitivity and specificity of the ELISA kits. The given diagnostic sensitivity and specificity of IgG ELISA was 96.6% and 98.2%, respectively, and that of IgM ELISA was be 95.8% and 100.0%, respectively. Apparent and true seroprevalence were calculated using Epi Tools (Rogan and Gladen, 1978; Brown et al., 2001; Sergeant, 2018).

### Descriptive and univariable analysis

Descriptive, univariable and multivariable analyses were conducted in R statistical program (R statistical package version 3.4.0, R Development Core Team [2015], http://www.r-project.org).

Descriptive analyses of all the variables were performed. The continuous variables ‘age’ and ‘years in veterinary practice’ were not normally distributed, and hence were categorised by using their quartiles. Fisher’s exact test (one tailed) was conducted when assumptions of Chi– squared test were not met. Variables in the univariable analyses having a likelihood ratio chi– square p–value of <0.25 and having less than 10% missing values were included for multivariable model building.

### Multivariable analyses

We constructed a mixed effects logistic regression model using forward selection (likelihood ratio), stepwise approach. The variable ‘region’ was used as a random effect to account for clustering at this level. The explanatory variables having a p-value of <0.05 were retained and those having p-value >0.25 in the univariable analysis were tested in the final model. Generalized variable inflation factor (GVIF) was estimated to determine multicollinearity in the final model and the variables having GVIF^(1/(2 *Df)) more than 2 were eliminated. Variables in the final model were tested for biologically important two-way interactions. Model adequacy was determined using the likelihood ratio chi-squared goodness-of-fit statistic and residuals.

## Results

### Apparent and estimated true seroprevalence

The apparent and estimated true seroprevalence of *T. gondii* antibodies using IgG ELISA was found to be 8.78% (95% CI: 5.63%–13.45%) and 7.36% (95% CI: 4.04%–12.29%), respectively (Table 1). The apparent and estimated true seroprevalence using IgM ELISA was found to be 0.49% (95% CI: Inestimable–2.71%) and 0.51% (95% CI: Inestimable– 2.83%), respectively.

Frequency distributions of the 31 categorical explanatory variables have been reported in Table S1. Overall, 205 veterinary personnel participated in this study. There were 10.2% academicians, 30.2% veterinary doctors, 17.6% veterinary pharmacists, 29.7% veterinary students and 12.2% animal attendants/class IV employees. Most of the participants reside in urban (71.2%) as compared to rural (28.8%) areas. Most of the participants were running mixed (handling both small and large animals – 87.32%) practice. More than 30% of the participants handled cat cases in past three months and approximately 6.0% of the participants contacted cat faeces in the last three months.

### Univariable results

Univariable results for the explanatory variables with the outcome variable having a p–value of less than 0.25 are presented in Table 2. Information for the remaining explanatory variables is presented in Supplementary appendix. Consuming mutton (p=0.029) and type of treatment of drinking water (p=0.017) were strongly associated with the outcome variable.

**Table 2.**
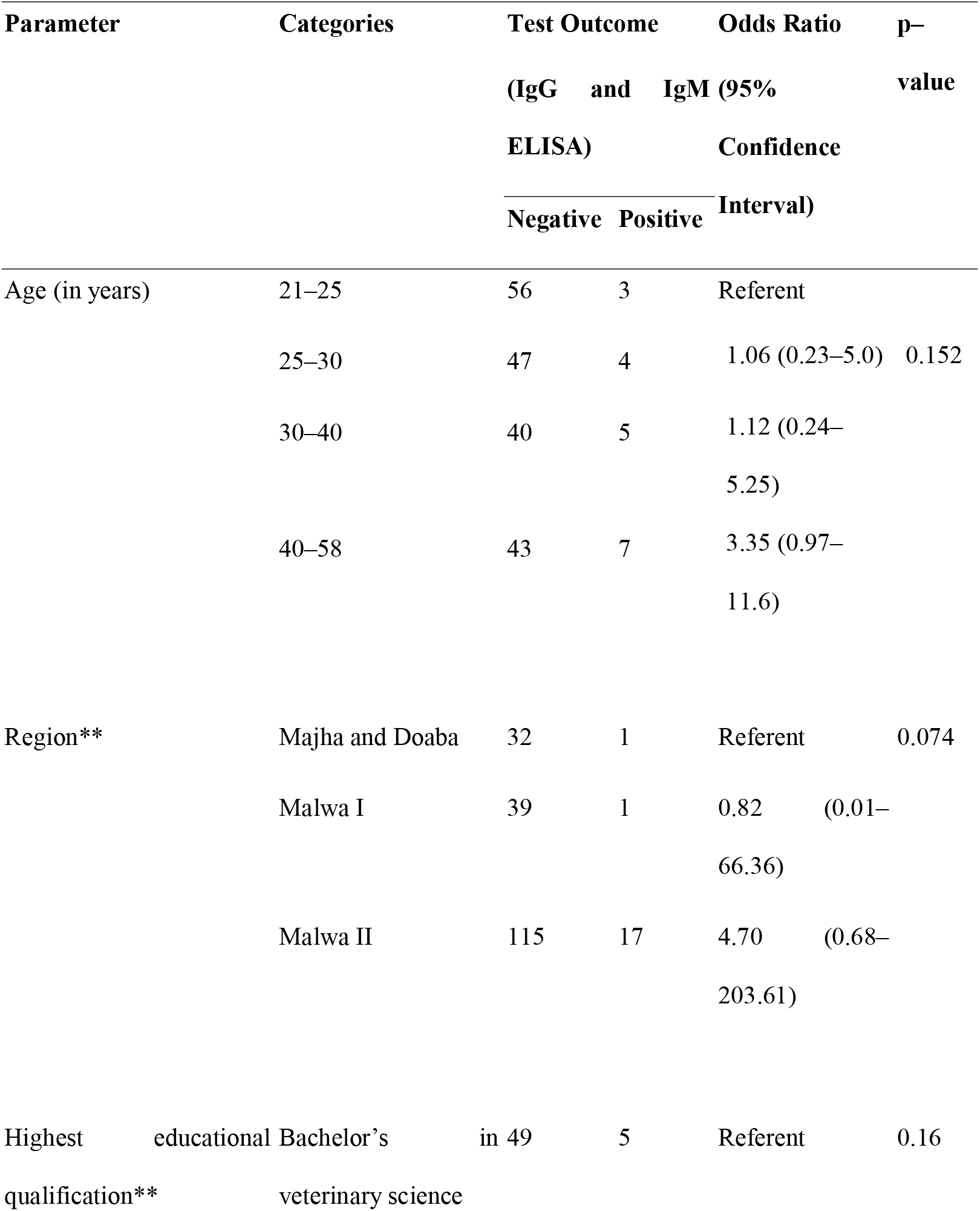
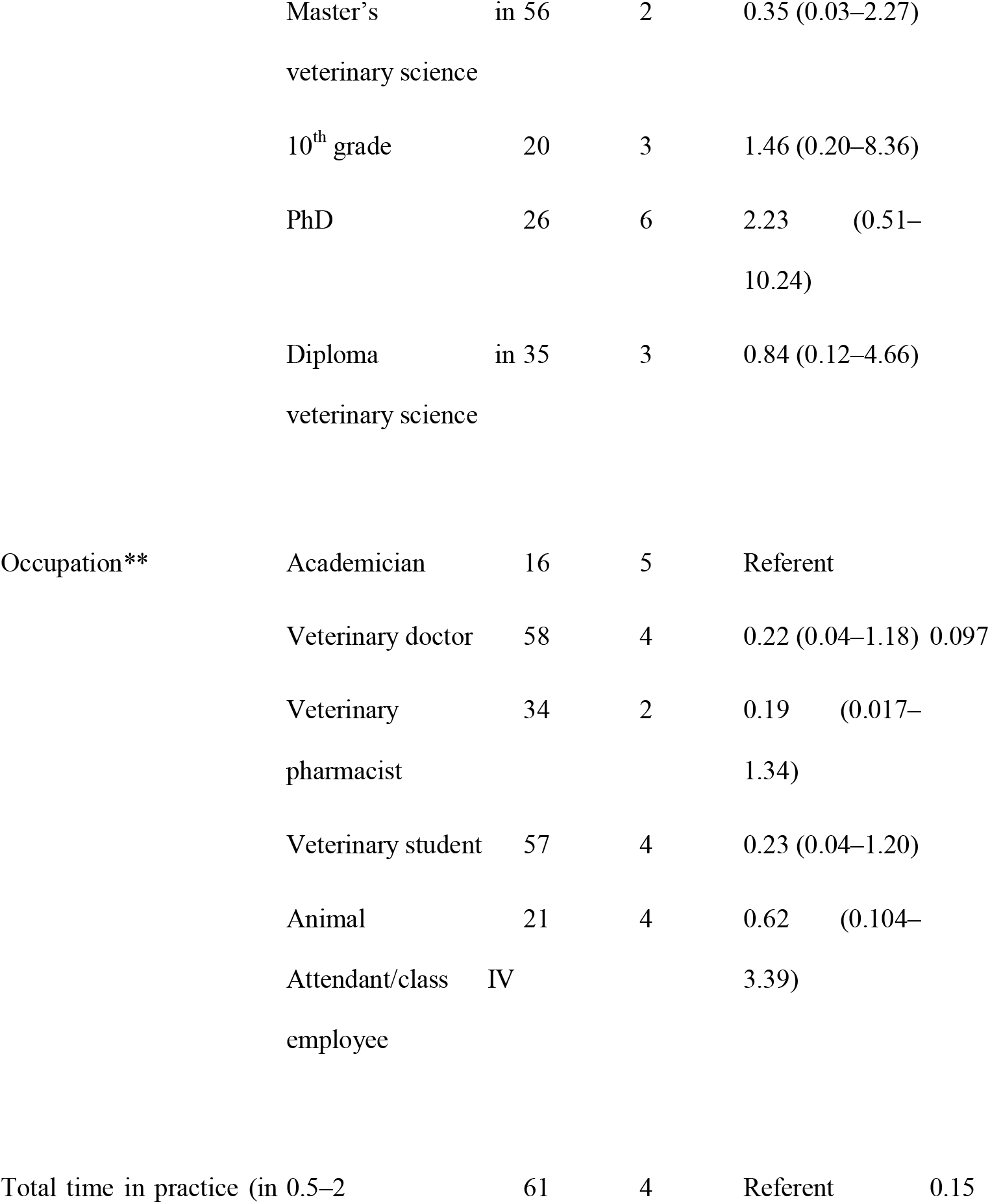
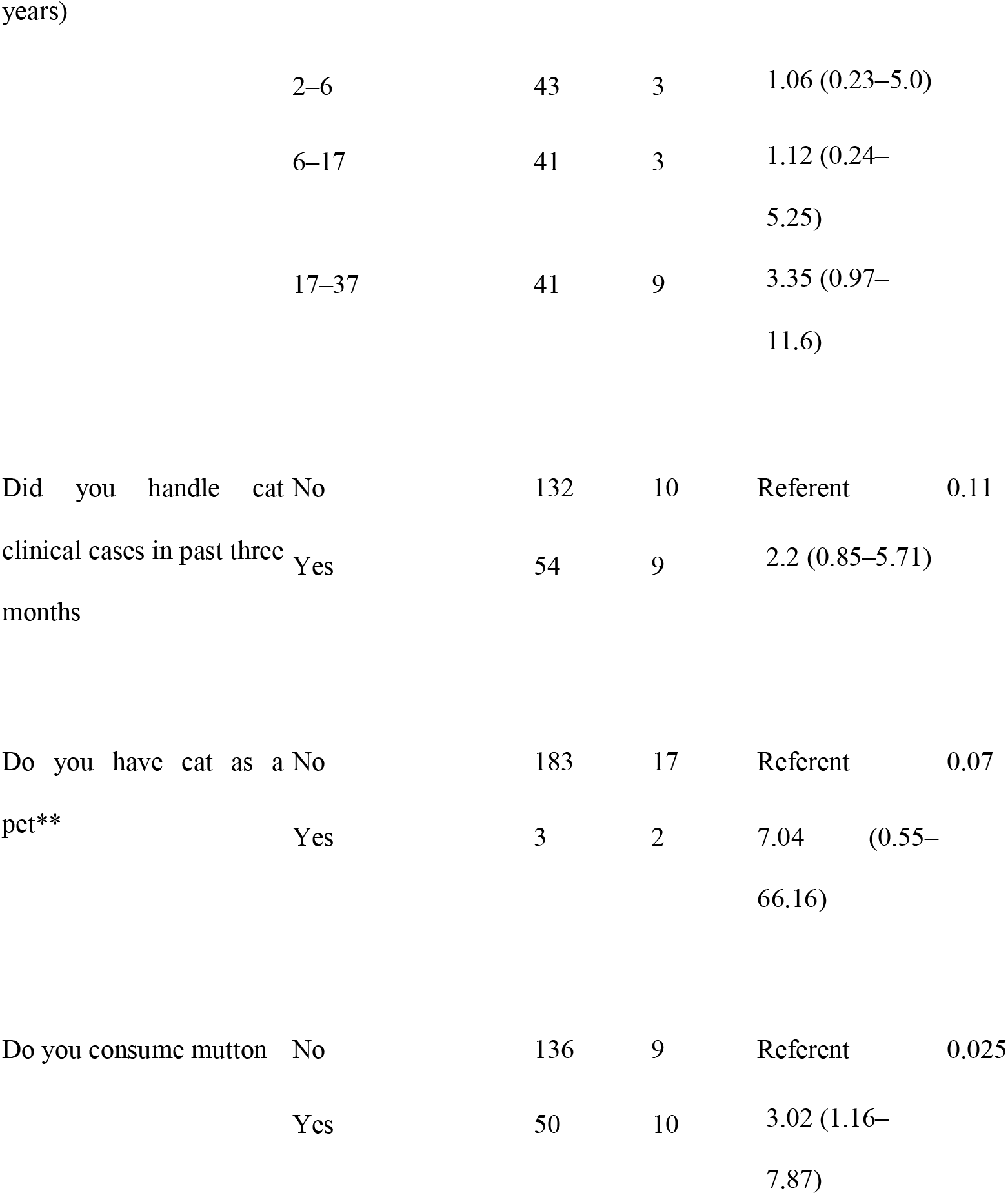
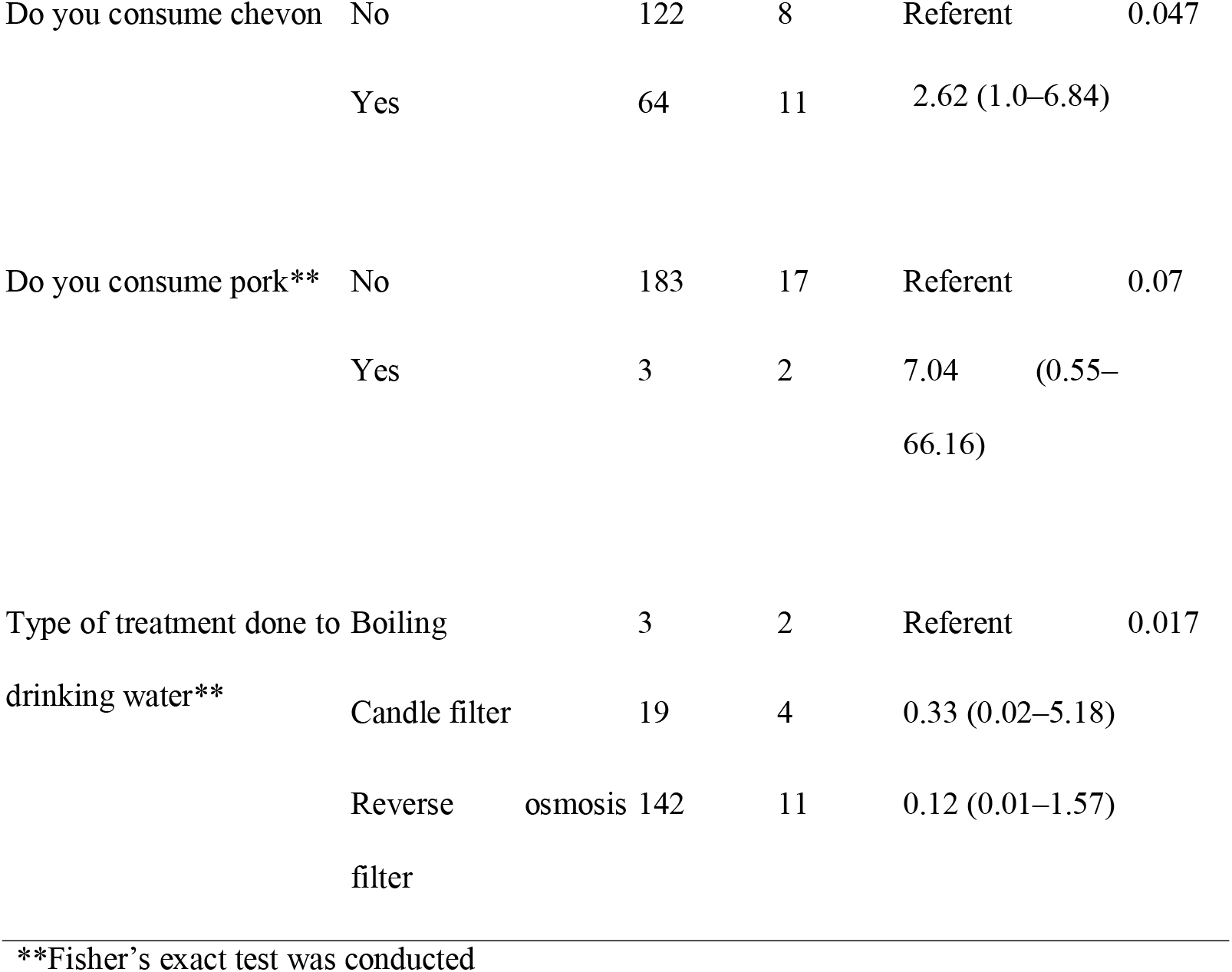
Contingency tables and univariable results (variables having p=<0.25) for *Toxoplasma* seropositivity (IgG and IgM ELISA) in a study of 205 veterinary personnel conducted in India in 2017–18.

### Multivariable results

The final multivariable model for *Toxoplasma* seropositive is presented in Table 3. After adjusting for other variables in the final model, consuming sheep meat and keeping cat as a pet were associated with large odds of having a positive test (Table 3).

**Table 3.**
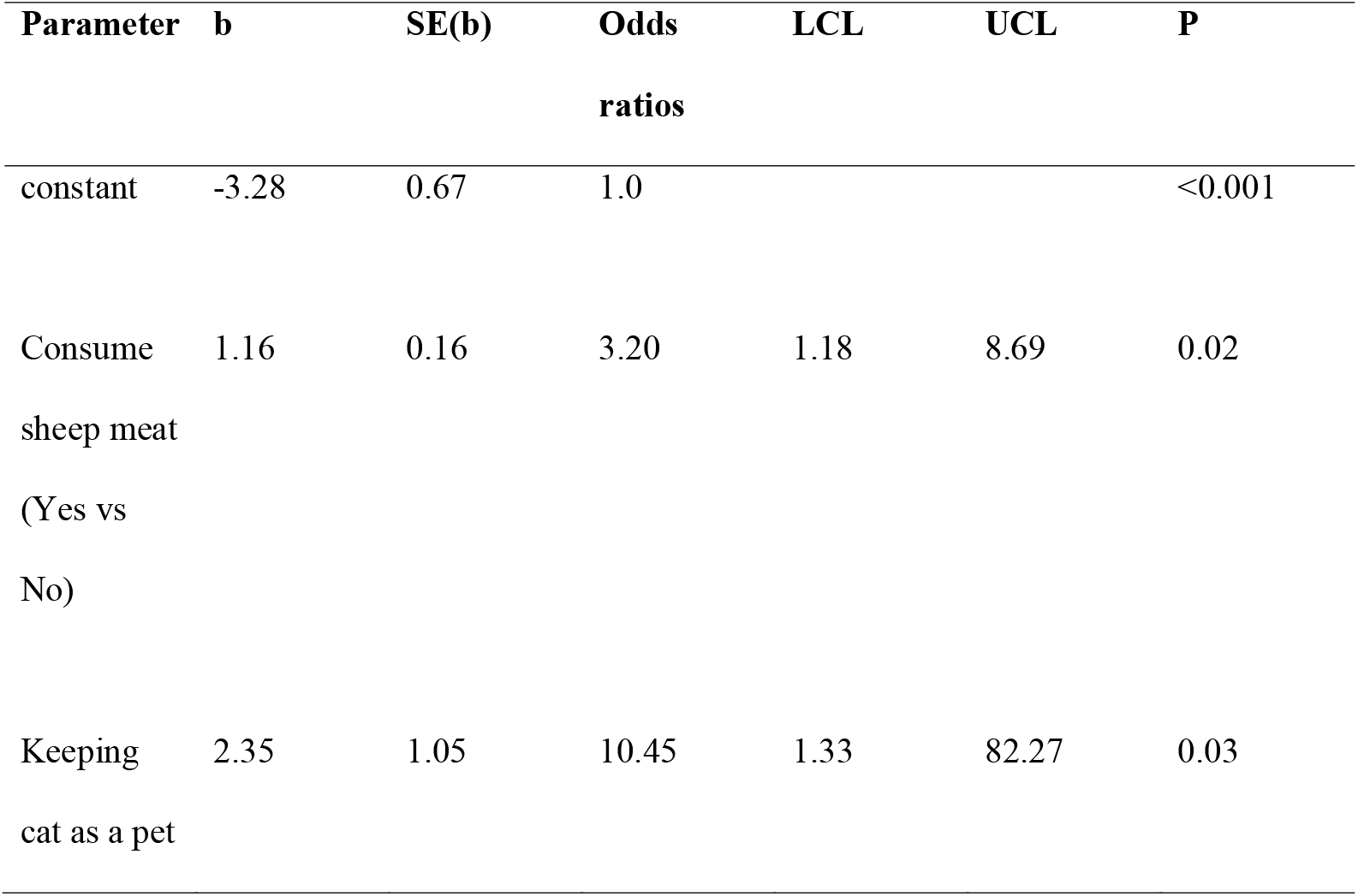
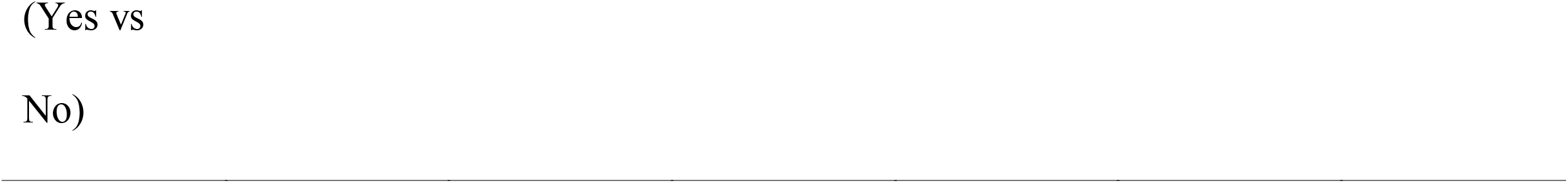
Final multivariable model for *Toxoplasma* seropositive outcome in a study of 205 veterinary personnel and students conducted in Punjab, India in 2017–18.

| Parameter | b | SE(b) | Odds<br>ratios | LCL | UCL | P |
| --- | --- | --- | --- | --- | --- | --- |
| constant | -3.28 | 0.67 | 1.0 |  |  | <0.001 |
| Consume<br>sheep meat<br>(Yes vs<br>No) | 1.16 | 0.16 | 3.20 | 1.18 | 8.69 | 0.02 |
| Keeping<br>cat as a pet | 2.35 | 1.05 | 10.45 | 1.33 | 82.27 | 0.03 |

## Discussion

This is the first study conducted systematically to determine seroprevalence and risk factor investigation for the exposure of *T. gondii* among veterinary personnel in north India. As far as we are aware, no studies to capture occupational exposure in veterinary personnel except one from western India (reporting 15% *Toxoplasma* seropositivity in zoo workers) have been conducted (RG et al., 2006). We believe such studies are required to aware and safeguard veterinary workforce from occupationally acquired diseases.

We estimated apparent seroprevalence of *T. gondii* antibodies using IgG ELISA to be 8.78% (95% CI: 5.63%–13.45%) in veterinary personnel in Punjab, India. Previous studies report presence of anti-*Toxoplasma gondii* antibodies (IgG or IgM) in 5% to 80% human populations in India (Khan et al., 2017). Lower seroprevalence estimates might be due to the fact that most of the veterinary personnel and students are well-aware of transmission risk of *Toxoplasma* and maintain high personal hygiene. In addition, significantly drier conditions in this region lower the survival of *T. gondii* oocysts (Dhumne et al., 2007).

We report that occupational exposure does not play an important role for the exposure of toxoplasmosis in veterinary personnel in Punjab state of India. Most of the known occupational risk factors such as type of practice, handling felines and cat faeces, and handling abortion cases were not significant in this study. However, this could not be absolutely ruled out. The single individual was IgM seropositive; he had no pet cat but had been handling abortion cases. We believe that this needs to be further investigated. Similarly, Sadaghian and Jafari (Sadaghian and Jafari, 2016) reported that toxoplasmosis is not occupationally related to veterinary laboratory science students. On the other hand, several studies have reported significant occupational risk of toxoplasmosis (Siponen et al., 2019). For example, occupational risks such as veterinarians living in rural areas, and not doing small animal practice were reported to be associated with *Toxoplasma* seropositive status in Finnish veterinary professionals (Siponen et al., 2019).

We found non-occupational risk factors e.g keeping cat as a pet and consuming sheep meat to be significantly associated with *Toxoplasma* seropositive status. Similarly, several non-occupational risk factors have been identified in the transmission of *T. gondii*. Jones and colleagues (Jones et al., 2009) identified important risks for the exposure to *Toxoplasma* including owning cats, and eating raw or uncooked pork, lamb, mutton, beef, game or mincemeat products to recently infected persons in Brazil. Dong and colleagues (Dong et al., 2018) also reported eating raw or undercooked meat and low level of risk to be important factors for toxoplasmosis. However, they also reported butchers, animal traffickers and zookeepers at a higher risk for toxoplasmosis.

Our study had certain limitations. We could not compare disease prevalence and risk with general public and other occupations such as meat handlers and cat owners. In addition, we have not tested veterinary personnel from other districts of the state. However, we believe that similar livestock production and animal husbandry practices exist across the state and this will not affect the overall results of this study.

Overall, we report a low seroprevalence of toxoplasmosis as compared to previous studies conducted on general public or non-occupationally exposed persons in India. The results reassure veterinary personnel that their occupational exposure does not enhance risk of getting infected with *T. gondii*. However, owning cat and consuming mutton are related with *Toxoplasma* seropositivity in Punjab, India.

## Data Availability

Data is available on request from the authors

