## Supplementary appendix for "Seroprevalence and risk factor investigation for the exposure of *Toxoplasma gondii* among veterinary personnel in Punjab, India"

Table S1. Frequency tables for 31 categorical explanatory variables in a risk factor study for toxoplasmosis among 205 veterinary personnel and students in Punjab, India in 2017–18.

| **Variable** | **Category** | **Frequency** | **Relative Frequency (%)** |
| --- | --- | --- | --- |
| **Demography** | | | |
| Region | Majha+Doaba | 33 | 16.09 |
|  | Malwa I | 40 | 19.52 |
|  | Malwa II | 132 | 64.39 |
| Residential area | Rural | 59 | 28.8 |
|  | Urban | 146 | 71.2 |
| Age (in years) | 21–25 | 59 | 28.8 |
|  | 25–30 | 51 | 24.9 |
|  | 30–40 | 45 | 21.9 |
|  | 40–58 | 50 | 24.4 |
| Gender | Female | 48 | 23.4 |
|  | Male | 157 | 76.6 |
| Highest educational qualification | B.V.Sc | 54 | 26.3 |
|  | M.V.Sc | 58 | 28.3 |
|  | Matriculation | 23 | 11.2 |
|  | Ph.D. | 32 | 15.6 |
|  | Vet Diploma | 38 | 18.5 |
| Occupation | Academicians | 21 | 10.24 |
|  | Veterinary Doctor | 62 | 30.24 |
|  | Veterinary Pharmacist | 36 | 17.56 |
|  | Veterinary Student | 61 | 29.76 |
|  | Animal Attendants/class IV  employees | 25 | 12.19 |
| Time in practice (in years) | 0.5–2 | 65 | 31.8 |
|  | 2–6 | 46 | 22.4 |
|  | 6–17 | 44 | 21.4 |
|  | 17–37 | 50 | 24.4 |
| **Occupational exposure** | | | |
| Type of practice | Large animals | 24 | 11.71 |
|  | Mixed | 179 | 87.32 |
|  | Small animals | 2 | 0.98 |
| Do you handle equines | No | 101 | 49.3 |
|  | Yes | 104 | 50.7 |
| Do you handle canines | No | 28 | 13.7 |
|  | Yes | 177 | 86.3 |
| Do you handle felines | No | 117 | 57.1 |
|  | Yes | 88 | 42.9 |
| Do you handle caprine | No | 80 | 39.0 |
|  | Yes | 125 | 61.0 |
| Do you handle porcine | No | 145 | 70.7 |
|  | Yes | 60 | 29.3 |
| Do you handle ovine | No | 109 | 53.2 |
|  | Yes | 96 | 46.8 |
| Have you handled cat cases in past three months | No | 142 | 69.3 |
|  | Yes | 63 | 30.7 |
| Have you handled cat faeces in past three months | No | 192 | 93.7 |
|  | Yes | 13 | 6.3 |
| If handled cat faeces, with or without gloves | No | 10 | 76.9 |
|  | Yes | 3 | 23.1 |
| Have you handled abortion cases in past three months | No | 82 | 40.0 |
|  | Yes | 123 | 60.0 |
| If you handled abortion cases, with gloves | No | 25 | 20.3 |
|  | Yes | 98 | 79.7 |
| If you handled abortion cases, with apron | No | 57 | 46.3 |
|  | Yes | 66 | 53.7 |
| **Non–occupational exposure** | | | |
| Do you have cat as pet | No | 200 | 97.6 |
|  | Yes | 5 | 2.4 |
| Are you vegetarian or non–vegetarian | Vegetarian | 136 | 66.3 |
|  | Non–vegetarian | 69 | 33.7 |
| Do you consume mutton | No | 76 | 55.9 |
|  | Yes | 60 | 44.1 |
| Do you consume chevon | No | 61 | 44.9 |
|  | Yes | 75 | 55.1 |
| Do you consume pork | No | 131 | 96.3 |
|  | Yes | 5 | 3.7 |
| Do you consume raw vegetables and fruits | No | 14 | 6.8 |
|  | Yes | 191 | 93.2 |
| What is the source of your portable water | Hand pump | 3 | 1.5 |
|  | Tap water | 202 | 98.5 |
| Do you treat drinking water | No | 24 | 11.7 |
|  | Yes | 181 | 88.3 |
| What type of treatment you do to your drinking water | Boiling | 5 | 2.7 |
|  | Candle filter | 23 | 12.7 |
|  | R.O. | 153 | 84.6 |
| Do you do gardening | No | 140 | 68.3 |
|  | Yes | 65 | 31.7 |
| Do you do garden with or without gloves | with gloves | 3 | 4.6 |
|  | without gloves | 62 | 95.4 |

Table S2. Contingency tables and univariable results (variables having p=>0.25) for *Toxoplasma* seropositivity (IgG and IgM ELISA) in a study of 205 veterinary personnel conducted in India in 2017–18.

| **Parameter** | **Categories** | **Test Outcome**  **(IgG and IgM ELISA)** | | **Odds Ratio**  **(95% Confidence Interval)** | **p–value** |
| --- | --- | --- | --- | --- | --- |
|  |  | **Negative** | **Positive** |  |  |
| Residential area | Rural | 53 | 6 | 0.86 (0.31–2.39) | 0.78 |
|  | Urban | 133 | 13 | Referent |  |
| Sex** | Female | 43 | 5 | Referent | 0.78 |
|  | Male | 143 | 14 | 0.84 (0.26–3.16) |  |
| Type of practice** | Large | 21 | 3 | Referent | 0.57 |
|  | Mixed | 163 | 16 | 0.68 (0.17–3.99) |  |
|  | Small | 2 | 0 | 0.0 (0.0–46.97) |  |
| Do you handle equines | No | 94 | 7 | Referent | 0.253 |
|  | Yes | 92 | 12 | 1.75 (0.66–4.65) |  |
| Do you handle canines** | No | 24 | 4 | Referent | 0.30 |
|  | Yes | 162 | 15 | 0.56 (0.16–2.50) |  |
| Do you handle felines | No | 107 | 10 | Referent | 0.682 |
|  | Yes | 79 | 9 | 1.22 (0.47–3.14) |  |
| Do you handle caprine | No | 71 | 9 | Referent | 0.438 |
|  | Yes | 115 | 10 | 0.69 (0.27–1.77) |  |
| Do you handle porcine | No | 131 | 14 | Referent | 0.764 |
|  | Yes | 55 | 5 | 0.85 (0.29–2.48) |  |
| Do you handle ovine | No | 99 | 10 | Referent | 0.961 |
|  | Yes | 87 | 9 | 1.02 (0.4–2.64) |  |
| Have you handled cat faeces in past three months** | No | 175 | 17 | Referent | 0.34 |
|  | Yes | 11 | 2 | 1.84 (0.18–9.68) |  |
| Handled cat faeces with or without gloves** | With gloves | 9 | 1 | Referent | 0.359 |
|  | Without gloves | 2 | 1 | 4.5 (0.19–106.81) |  |
| Have you handled abortion case in past three months | No | 75 | 7 | Referent | 0.767 |
|  | Yes | 111 | 12 | 1.16 (0.44–3.08) |  |
| Have you handled abortion case with gloves** | No | 23 | 2 | Referent | 1.0 |
|  | Yes | 88 | 10 | 1.30 (0.25–13.06) |  |
| Have you handled abortion case wearing an apron | No | 50 | 7 | Referent | 0.381 |
|  | Yes | 61 | 5 | 0.59 (0.18–1.96) |  |
| Are you vegetarian or non–vegetarian | Non–vegetarian | 122 | 14 | Referent | 0.469 |
|  | Vegetarian | 64 | 5 | 0.68 (0.23–1.97) |  |
| Do you consume raw vegetables and fruits** | No | 12 | 2 | Referent | 0.62 |
|  | Yes | 174 | 17 | 0.58 (0.11–5.84) |  |
| What is the source of your portable water** | Hand pump | 3 | 0 | Referent | 1.0 |
|  | Tap water | 183 | 19 | Inestimable (0.04–inestimable) |  |
| Do you treat drinking water** | No | 22 | 2 | Referent | 0.865 |
|  | Yes | 164 | 17 | 1.14 (0.25–5.27) |  |
| Do you do gardening | No | 129 | 11 | Referent | 0.317 |
|  | Yes | 57 | 8 | 1.65 (0.63–4.31) |  |
| Do you do garden with or without gloves** | with gloves | 2 | 1 | Referent | 0.33 |
|  | without gloves | 55 | 7 | 0.26 (0.012–17.14) |  |

**Fisher’s exact test was conducted
